# Effect of Blood Type on Mortality Among Patients with Brain Metastases

**DOI:** 10.1101/2022.11.22.22282634

**Authors:** Yilong Zheng, Jiashen Zhao, Kejia Teo, Vincent Diong Weng Nga, Tseng Tsai Yeo, Mervyn Jun Rui Lim

## Abstract

**Objective:** ABO blood type has been associated with mortality among patients with cancer, but this association has thus far not been investigated among patients with brain metastases. Hence, we aimed to investigate the association between ABO blood type and mortality among patients who underwent surgical resection of brain metastases.

**Methods:** A single-center retrospective study of patients who underwent surgical resection of brain metastases between 2011 and 2019 was conducted. Cox proportional hazards models were constructed, adjusting for potential confounders, to evaluate whether blood type was independently associated with overall mortality.

**Results:** A total of 158 patients were included in the analysis. The mean (SD) age of the cohort was 59.3 (12.0) years, and 67.7% of patients were female. The median overall survival of patients with AB was 11.2 months, while the median overall survival of patients with blood types O, B, and A were 11.7, 13.5, and 14.4 months respectively. On multivariate analysis adjusting for potential confounders, there was a statistically significant association between blood type AB and a higher risk of overall mortality (HR=2.42, 95% CI=1.20, 4.86, p=0.013).

**Conclusions:** Blood type AB was independently associated with a higher risk of overall mortality among patients who underwent surgical resection of brain metastases. We hypothesize that antibodies against blood type antigens A and/or B have anti-tumor activity in patients with brain metastases. Further studies are needed to validate our conclusions and hypothesis.

## Introduction

Brain metastases are the commonest type of malignant tumor to affect the central nervous system.^1-3^ ABO blood type has been associated with mortality among patients with cancer, including but not limited to cancers of the lung,^4, 5^ bladder,^6^ pancreas,^7, 8^ larynx,^9^ and liver,^10^ but the specific blood type that was associated with mortality was inconsistent.^4-10^ There were also studies which found no statistically significant association between blood type and mortality among patients with cancer.^11-13^ Proposed mechanisms for the association between ABO blood type and mortality include the influence of ABO blood type on the body’s inflammatory state and therefore the body’s response to the tumor.^4, 5^

The association between ABO blood type and mortality among patients with brain metastases has thus far not been studied. Therefore, we aimed to investigate the association between ABO blood type and mortality in our cohort of patients who underwent surgical resection of brain metastases.

## Methods

### Study design

This was a retrospective study of patients who underwent surgical resection of brain metastases at our institution between March 2011 and December 2019. The operating theatre records database was accessed to retrieve the National Registration Identity Card numbers (NRICs, which act as the national identification number) of patients who underwent surgical resection of a brain tumor. The medical records of the patients were then screened for inclusion in the study. The inclusion criteria were (a) patients who had a histologically verified metastasis to the brain, and (b) patients who were 18 years old and above on the day of surgery. Institutional ethics approval was obtained from the local institutional review board prior to the commencement of the study (National Healthcare Group Domain Specific Review Board; Reference Number 2020/00358), and a waiver of informed consent was granted since this study posed no more than minimal risk to participants.

### Data collection

Clinical data were collected using a standardized data collection form. Variables collected include (1) demographics, including age on the day of surgical resection, sex (defined as male or female), and ethnicity (defined as Chinese, Malay, Indian, or Others), (2) results of the preoperative full blood count, including hemoglobin level, mean corpuscular volume, hematocrit, white blood cell, red blood cell, platelet, neutrophil, lymphocyte, monocyte, eosinophil, and basophil count, (3) the blood type of the patients, defined as A, AB, B, and O, (4) disease factors, including the site of the primary tumor (defined as the 5 commonest sites and ‘others’), whether there were extracranial metastases preoperatively, volume of the largest tumor on preoperative brain MRI (as approximated by the ABC/2 formula, where A, B, and C denote the largest antero-posterior, medial-lateral, and cranio-caudal diameters of the lesion respectively), number of tumors on the preoperative brain MRI, (5) whether the patient passed away, and (6) the dates of the first surgical resection of the brain metastases, latest follow-up, and demise (if applicable).

### Statistical analysis

The baseline characteristics of the patients were reported using mean and standard deviation for continuous variables and count numbers and percentages for categorical variables. Hypothesis testing was conducted using the chi-square test for categorical data, and the student’s t-test for continuous data. Hypothesis testing was conducted using Fisher’s exact test for variables with any category that contained less than 5 observations. A p-value of 0.050 or less was taken to be statistically significant.

To ascertain whether blood type was independently associated with overall mortality, Cox regression models were constructed with blood type as the exposure, adjusting for age, sex, ethnicity, site of the primary tumor, and variables that had a statistically significant association with blood type on univariate analysis. A p-value of 0.050 or less was taken to be statistically significant. Time-to-mortality was defined as the duration between the first surgical resection and the date of the latest follow-up. All data analyses were conducted using R Studio Version 1.2.5042.

## Results

### Baseline characteristics of the study population

The baseline characteristics of the study population were reported in Table 1. A total of 158 patients were included in the analysis. The mean (SD) age of the study population was 59.3 (12.0) years, and 67.7% of patients were female. The commonest blood type in our cohort was O (55 patients, 34.8%), followed by A (46 patients, 29.1%), B (43 patients, 27.2%), and AB (14 patients, 8.9%).

**Table 1:** Baseline characteristics of patients who underwent surgical resection of brain metastases stratified by blood group

| Variable | O (n=55,<br>34.8%) | A (n=46,<br>29.1%) | B (n=43,<br>27.2%) | AB (n=14,<br>8.9%) | Total<br>(n=158) | p-value |
| --- | --- | --- | --- | --- | --- | --- |
| Age; mean (SD) | 60.2 (11.2) | 60.4 (11.6) | 57.5 (12.1) | 57.9 (15.8) | 59.3 (12.0) | 0.602 |
| Ethnicity; *n* (%) |  |  |  |  |  | 0.997 |
| Chinese | 37 (67.3) | 30 (65.2) | 30 (69.8) | 10 (71.4) | 107 (67.7) |  |
| Malay | 8 (14.5) | 7 (16.3) | 7 (15.2) | 2 (14.3) | 24 (15.2) |  |
| Indian | 3 (5.5) | 2 (4.3) | 2 (4.7) | 0 (0.0) | 7 (4.4) |  |
| Others | 7 (15.2) | 7 (15.2) | 4 (9.3) | 2 (14.3) | 20 (12.7) |  |
| Female; *n* (%) | 23 (41.8) | 29 (63.0) | 26 (60.5) | 6 (42.9) | 84 (53.2) | 0.104 |
| Site of primary tumor;<br>*n* (%) |  |  |  |  |  | 0.898 |
| Lung | 24 (43.6) | 19 (41.3) | 16 (37.2) | 4 (28.6) | 63 (39.9) |  |
| Breast | 15 (27.3) | 8 (17.4) | 14 (32.6) | 3 (21.4) | 40 (25.3) |  |
| Colorectum | 6 (10.9) | 8 (17.4) | 2 (4.7) | 2 (14.3) | 18 (11.4) |  |
| Skin | 2 (3.6) | 2 (4.3) | 2 (4.7) | 1 (7.1) | 7 (4.4) |  |
| Kidney | 2 (3.6) | 2 (4.3) | 1 (2.3) | 1 (7.1) | 6 (3.8) |  |
| Others | 6 (10.9) | 7 (15.2) | 8 (18.6) | 3 (21.4) | 24 (15.2) |  |
| Presence of extracranial<br>metastases<br>preoperatively; *n* (%) | 29 (52.7) | 33 (71.7) | 29 (67.4) | 9 (64.3) | 100 (63.3) | 0.223 |
| Number of tumors on<br>preoperative brain<br>MRI; mean (SD) | 2.9 (6.7) | 2.5 (2.1) | 2.5 (2.1) | 2.4 (2.3) | 2.6 (4.3) | 0.953 |
| Volume of largest<br>tumor on preoperative<br>brain MRI (cm<sup>3</sup>); mean<br>(SD) | 22.1 (37.2) | 22.6 (43.0) | 20.2 (22.8) | 21.3 (20.2) | 21.7 (34.5) | 0.990 |
| Full blood count*; *n*<br>(%) |  |  |  |  |  |  |
| White blood cell count | 9.3 (3.0) | 10.1 (4.2) | 11 (3.7) | 9.9 (3.5) | 10 (3.7) | 0.153 |
| Red blood cell count | 4.5 (0.8) | 4.1 (0.6) | 4.7 (0.6) | 4.4 (0.9) | 4.4 (0.7) | **0.002** |
| Hemoglobin | 12.9 (1.8) | 12.3 (1.8) | 13.2 (1.6) | 12.6 (2.2) | 12.8 (1.8) | 0.132 |
| MCV | 87.7 (7.5) | 89 (6.1) | 84.4 (8.4) | 87.1 (6.4) | 87.1 (7.4) | **0.024** |
| Hematocrit | 38.9 (4.5) | 36.6 (5.3) | 39.4 (4.3) | 37.9 (6.8) | 38.3 (5.1) | **0.032** |
| Platelet count | 255.4 (83.6) | 254.7<br>(102.9) | 261.1 (63.5) | 231.3 (68.2) | 254.6 (83.5) | 0.719 |
| Neutrophil count | 8.1 (4.4) | 8.1 (4.2) | 10.3 (8.7) | 8.2 (3.4) | 8.7 (5.8) | 0.213 |
| Lymphocyte count | 1.3 (0.5) | 1.7 (2.0) | 1.4 (1.0) | 2 (3.5) | 1.5 (1.6) | 0.361 |
| Monocyte count | 0.6 (0.2) | 0.7 (0.4) | 0.5 (0.3) | 0.9 (1.6) | 0.6 (0.5) | 0.071 |
| Eosinophil count | 0.1 (0.1) | 0.1 (0.3) | 0.1 (0.1) | 0.1 (0.2) | 0.1 (0.2) | 0.724 |
| Basophil count | 0.0 (0.0) | 0.0 (0.0) | 0.0 (0.0) | 0.0 (0.0) | 0.0 (0.0) | 0.857 |

\*Count numbers reported are of patients with a normal value for the test.
Abbreviations: SD, standard deviation; MCV, mean corpuscular volume; MRI, magnetic resonance imaging.

On univariate analysis, there was a statistically significant association between blood type and preoperative red blood cell count (p=0.002), mean corpuscular volume (p=0.024), and hematocrit (p=0.032).

### Blood type and mortality

The median overall survival of patients with blood type AB was 11.2 months, while the median overall survival of patients with blood types O, A, and B were 11.7, 13.5, and 14.4 months respectively. The mean (SD) duration of follow-up was 15.5 (17.6) months.

On time-to-event analysis, there was a statistically significant association between blood type AB and overall mortality on both univariate analysis (HR=2.24, 95% CI=1.15, 4.38, p=0.018) and multivariate analysis adjusting for potential confounders (HR=2.42, 95% CI=1.20, 4.86, p=0.013) (Table 2).

**Table 2:** Cox regression model of blood type against overall mortality, adjusting for potential confounders

| Variable | Unadjusted HR (95% CI) | p-value | Adjusted HR (95% CI)<sup>1</sup> | p-value |
| --- | --- | --- | --- | --- |
| Blood type |  |  |  |  |
| A | — | — | — | — |
| AB | 2.24 (1.15, 4.38) | **0.018** | 2.42 (1.20, 4.86) | **0.013** |
| B | 1.08 (0.66, 1.76) | 0.760 | 1.07 (0.63, 1.84) | 0.795 |
| O | 1.20 (0.76, 1.89) | 0.443 | 1.29 (0.81, 2.06) | 0.288 |

<sup>1</sup>Adjusted for age, sex, ethnicity, site of primary cancer, and variables that had a statistically significant association with blood type on univariate analysis, including red blood cell count, mean corpuscular volume, and hematocrit.
Abbreviations: HR, hazard ratios; CI, confidence intervals.

## Discussion

In our cohort of patients who underwent surgical resection of brain metastases, patients with blood type AB had a significantly higher risk of overall mortality. Data on the association between blood type and mortality among patients with cancer are conflicting, with different studies reporting statistically significant associations between different blood types and mortality.^4-10^ There were also studies that found no statistically significant association between blood type and mortality among patients with cancer.^11-13^

The association between blood type and mortality may be related to the different degrees of immune response to the tumor across different blood types. Li et al. reported that in their cohort of patients with curatively resected non-small cell lung cancer, patients with blood type AB had the lowest overall and disease-free survival rates.^5^ The authors attributed the findings to the influence of ABO blood type on the host inflammatory state.^5^ Specifically, the authors stated that single nucleotide polymorphisms at the ABO gene locus were associated with differing levels of circulating tumor necrosis factor-alpha (TNF-α),^14^ E-selectin,^15^ soluble intracellular adhesion molecule-1 (sICAM-1),^16, 17^ and P-selectin,^16^ all of which have been reported to be associated with inflammatory responses that were associated with angiogenesis, tumor growth, and tumor progression.^5^

On top of the differing levels of inflammatory mediators across different blood types, the independent association between blood type and mortality may also be due to the anti-tumor effect of antibodies against blood type antigens A and B. This is because in our cohort, patients with blood type AB had a significantly higher risk of mortality, and patients with blood type AB have no antibodies against blood type antigens A and B. On the other hand, patients with non-AB blood type had a significantly lower risk of mortality, and patients with non-AB blood type also have antibodies against blood type antigens A and/or B. Therefore, we hypothesize that antibodies against blood type antigens A and/or B have anti-tumor effect.

Our study has several limitations. First, the relatively small sample size of our cohort limited the statistical power of our analysis. Second, as our study included only patients from a single institution, the results may not be generalizable to other populations. Lastly, we did not collect and analyze data on the cause of death due to the incompleteness of documentation on the cause of death in the electronic medical records.

## Conclusion

Blood type AB was independently associated with a higher risk of overall mortality among patients who underwent surgical resection of brain metastases. We hypothesize that antibodies against blood type antigens A and/or B have anti-tumor activity in patients with brain metastases. Further studies are needed to validate our conclusions and hypothesis.

## Data Availability

All data produced in the present study are available upon reasonable request to the authors

## Acknowledgements

We thank Dr Yap Eng Soo and Dr Chan Wee Lee William for helpful comments on an initial draft of the manuscript.

